# Relationships between freezing of gait severity and cognitive deficits in Parkinson’s disease

**DOI:** 10.1101/2021.04.29.21256338

**Authors:** Jamie L. Scholl, Arturo I. Espinoza, Wijdan Rai, Matt Leedom, Lee A. Baugh, Patti Berg-Poppe, Arun Singh

**Author notes:** **Correspondence to:** Dr. Arun Singh, Division of Basic Biological Sciences, Sanford School of Medicine, University of South Dakota, 414 E. Clark St. Vermillion, SD, 57069, USA.

## Abstract

**Objective:** Evidence supports an association between freezing of gait (FOG) severity and cognitive functioning in patients with Parkinson’s disease (PD); however, results are varied. Here we sought to explore the differences in cognitive measurements via multivariable statistical models in patients with PD.

**Methods:** PD patients with (PDFOG+, n=41) and without FOG (PDFOG–, n=39) and control healthy subjects (n=41) participated in the study. The NIH toolbox cognition battery, Montreal cognitive assessment (MoCA), and interval timing task were used to test cognitive domains. Measurements were compared between groups using multivariable models and adjusting for covariates. Correlation analyses, linear regression, and mediation models were applied to examine relationships among disease duration and severity, FOG severity, and cognitive functioning.

**Results:** Significant differences were observed between controls and PD patients for all cognitive domains. PDFOG+ and PDFOG– exhibited differences in the dimensional change card sort (DCCS) test, interval timing task, and MoCA scores. After adjusting for covariates in two different models, PDFOG+ and PDFOG– differed in both MoCA and DCCS scores. In addition, significant relationships between FOG severity and cognitive function (MoCA, DCCS, and interval timing) were also found. Regression models suggest that FOG severity may be a predictor of cognitive impairment, and mediation models show the effects of cognitive impairment on the relationship between disease severity and FOG severity.

**Conclusions:** Overall, this study provides insight into the relationship between cognitive and FOG severity in patients with PD, which could aid in the development of therapeutic interventions to manage both.

## 1. Introduction

Freezing of gait (FOG) has attracted attention within clinical and scientific groups, as it is one of the most mismanaged motor symptoms in Parkinson’s disease (PD) [1, 2]. Approximately 80% of patients with advanced stage PD develop FOG, increasing risk of falls, loss of independence, reduced quality of life, and the expression of mood disorders [3]. FOG also occurs in ∼26% of patients in early-stage PD [4]; however, not all patients with PD go on to develop FOG symptoms, regardless of disease severity or duration. This variability suggests that different factors, such as age, depression, anxiety, sleeping problems; or physiological abnormalities, such as increased beta-band oscillations in the cortico-basal ganglia network, may be involved in the development of FOG in patients with PD [5, 6].

A recent study suggested the existence of PD patient subgroups exhibiting FOG (PDFOG+) on the basis of three predominant freezing triggers: motor type (occurrence of FOG during turn), cognitive type (occurrence of FOG during dual-task), or limbic type (occurrence of FOG during anxiety) [7]. A relationship between cognitive deficits and FOG in patients with PD has been proposed [8-10]; however, it remains undetermined whether cognitive impairment is the main contributor in FOG, or just one of the factors compounding FOG. Gait parameters (specifically, pace, variability, and postural control), rather than cognition, can be employed as a predictor for deterioration in cognitive domains in patients with early-stage PD [9]. Therefore, it is critical to further explore the relationship between FOG and cognitive function in PD.

Cognitive impairment is common in patients with PD as the disease advances. Previous studies have proposed that FOG can result from frontal malfunction or a disconnect between the frontal lobe and the basal ganglia [5, 6, 11, 12]. Recently, reduced power of midfrontal low-frequency oscillation has been shown during leg movements in PDFOG+ compared to PD patients who do not exhibit FOG (PDFOG–). As gait requires active cognitive and motor control systems [8, 12, 13], frontal theta- and beta-band oscillations in the cortico-basal ganglia network may be critical in initiating and executing gait and may also be involved in freezing among PD patients [6, 12]. Aside from physiological factors, studies have shown correlations between FOG and reduction in executive function, attention, memory, and visuospatial function, as well as with hallucinations [14, 15]. Though many previous studies did not account for age, disease severity, or disease duration, a recent study showed no significant difference in cognitive function between PDFOG+ and PDFOG– when adjusting for those covariates, particularly disease severity [10]. Aside from the previously discussed exploration into the one-sided relationship between cognitive dysfunction and FOG in patients with PD [11, 15], few studies have modeled the relationship between FOG and cognitive deficits [10, 16], or determined the directionality of this relationship.

The current study sought to further explore the differences in cognitive measurements between older healthy control participants, PDFOG–, and PDFOG+ via multivariable statistical models, including FOG severity, after adjusting for appropriate covariates. Regression analysis was also performed to determine if FOG severity could be used to predict cognitive impairment in patients with PD.

## 2. Materials and methods

### 2.1. Study participants

A total of 121 participants were recruited into the study: 81 patients with PD (41 PDFOG+ and 39 PDFOG–) and 41 healthy, age matched controls. All procedures were approved by the Institutional Review Boards at the University of Iowa and the University of South Dakota. Clinical demographics are shown in Table 1.

**Table 1.**
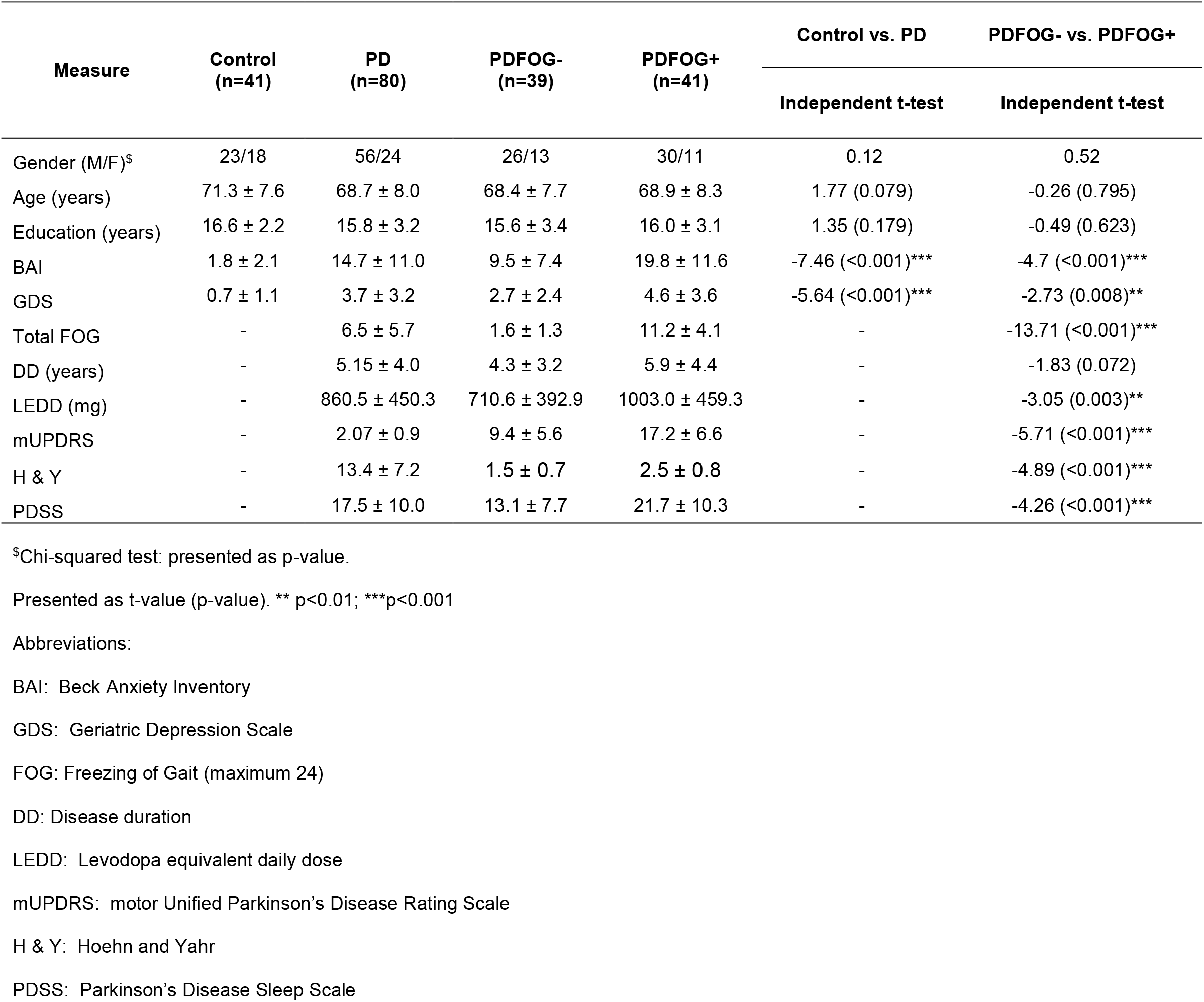
Demographic and clinical assessments in controls vs. patients with PD.

### 2.2. Identification of PDFOG+

Several criteria were implemented to identify and assign patients with PD to PDFOG+ or PDFOG– groups. Initially, patients confirmed they had difficulty in starting, stopping, and turning during a movement, required assistance while walking, or were wheelchair bound; and/or their score on the FOG questionnaire number 3 was greater than zero, suggestive of at least one episode of FOG in the past month [17]. Finally, an objective examination of the patient consisting of an unassisted walk with rapid turning was completed prior to study initiation. FOG episodes in PDFOG+ may cause fall risk during the off-medication state. Therefore, to match with real-word gait and cognitive functions, all PD participants were assessed with levodopa medication.

### 2.3. Clinical, motor, and cognitive assessments

Age, gender, and years of education were documented for each participant. Anxiety and depression were measured via the Beck Anxiety Inventory (BAI) and the Geriatric Depression Scale (GDS) for each subject. Sleep abnormality via the Parkinson’s Disease Sleep Scale (PDSS), disease duration in years, and levodopa equivalent daily dose were determined for patients with PD. To assess PD severity, the motor portion of the Unified Parkinson’s Disease Rating Scale (mUPDRS) [18] and the Hoehn and Yahr scale were determined. All clinical assessments were performed similar to our previous reports [12, 19].

A comprehensive cognitive function score was determined using the Montreal Cognitive Assessment (MoCA), which includes items measuring short term memory; visuospatial abilities; executive functions; attention, concentration, and working memory; language; and orientation to time and place. Five of seven instruments comprising the cognition battery of the NIH toolbox for assessment of neurological and behavioral function (NIHTB-CB) were used to compile scores for specific cognitive domains [20]. Episodic memory was measured using the NIHTB-CB Picture Sequence Memory (PSM) test. The NIHTB-CB Dimensional Change Card Sort (DCCS) and Flanker Inhibitory Control and Attention (FICA) instruments measured executive function and attention. Language domain results were captured using the NIHTB-CB Picture Vocabulary (PV) test, and the NIHTB-CB Pattern Comparison Processing Speed Test (PCPS) measured processing speed. Fully Corrected T-Scores, corrected for age and other demographic characteristics (education, gender, and race/ethnicity), were calculated using the NIH toolbox. A 7-second interval timing task was used to measure temporal performance for the continuous memory functioning index [19, 21]. The response time coefficient of variability (RTCV) was used for the analysis, as our previous study showed significant differences in RTCV between controls and PD patients, rather than mean response time [19].

### 2.4. Statistical analysis

Demographic and clinical assessments were compared between PD and controls and between PDFOG+ and PDFOG– using independent *t-tests* (gender with chi-squared test). Motor and cognitive measurements were first compared using independent *t-tests*, and then with ANCOVAs by applying analysis of variance for linear regression models while controlling for covariates. We used “fitlm” followed by “anova” Matlab (MathWorks, Inc) functions for ANCOVA models. Covariates of age, gender, and years of education, disease duration, as well as disease severity were selected for modeling. Spearman correlations and Spearman partial correlations were used to determine associations between cognitive measurements and FOG scores in patients with PD after controlling for covariates.

Furthermore, a fit linear regression model using “fitlm” function in Matlab was applied, followed by predictions for each observation using “predict” function to observe the relationship between FOG severity (as measured by FOG scores), cognitive function (as measured by MoCA scores), and disease severity (as measured by mUPDRS scores). Slopes of predicted values may determine the extent of the relationship between FOG severity and development of cognitive deficits. In addition, mediation analyses in “R” were performed to further determine if cognitive deficits mediated the relationship between disease severity and FOG severity. The mediate function in R gives average causal mediation effects (ACME), average direct effects (ADE), and combined indirect and direct effects (total effect). Statistical significance was set at α = 0.05. All values are shown in mean ± standard deviation.

## 3. Results

### 3.1. Differences in demographic and clinical assessments

There were no between-group differences in age, gender, and years of education; however, independent *t-tests* revealed differences between controls and patients with PD in both anxiety (controls, 1.8 ± 2.1; PD, 14.7 ± 11.0; p < 0.001) and depression (controls, 0.7 ± 1.1; PD, 3.7 ± 3.2; p < 0.001) measures, as well as when comparing PDFOG+ and PDFOG–. The PDFOG+ group exhibited significantly greater anxiety (19.8 ± 11.6) and depression scores (4.6 ± 3.6) compared to PDFOG– (9.5 ± 7.4, p < 0.001 and 2.7 ± 2.4 p = 0.008, respectively). When comparing PDFOG+ and PDFOG–, there were no differences in disease duration; however, differences were found in motor disease severity, medication, and sleep scores (Table 1).

### 3.2. Differences in cognitive measurements

Significant differences between control and patients with PD were found in all cognitive domains (Table 2). Patients with PD showed significantly worse performance in all cognitive tests even after adjusting for covariates of age, gender, and years of education (Table 2).

**Table 2.** Cognitive measurements in controls vs. patients with PD.

| Measure | Control<br>(n=41) | PD (n=80) | PDFOG-<br>(n=39) | PDFOG+<br>(n=41) | Control vs. PD |  | PDFOG- vs. PDFOG+ |  |  |
| --- | --- | --- | --- | --- | --- | --- | --- | --- | --- |
|  |  |  |  |  | Independent<br>t-test | <sup>a</sup>ANCOVA | Independent<br>t-test | <sup>a</sup>ANCOVA | <sup>b</sup>ANCOVA |
| *Cognitive function* |  |  |  |  |  |  |  |  |  |
| MoCA | 26.6 ± 1.8 | 24.3 ± 3.6 | 25.1 ± 3 | 23.5 ± 4 | 3.9<br>($<0.001$)*** | 14.53<br>($<0.001$)*** | 2.05<br>(0.044)* | 4.55<br>(0.034)* | 4.7<br>(0.034)* |
| *Episodic memory* |  |  |  |  |  |  |  |  |  |
| PSM | 55.5 ± 9.9 | 46.4 ± 11.7 | 48 ± 11.8 | 44.8 ± 11.6 | 4.11<br>($<0.001$)*** | 23.74<br>($<0.001$)*** | 1.2<br>(0.234) | 0.049<br>(0.487) | 0.43<br>(0.515) |
| *Executive function/Attention* |  |  |  |  |  |  |  |  |  |
| DCCS | 58.2 ± 13.2 | 50.1 ± 12.3 | 54.1 ± 12.2 | 46.1 ± 11.1 | 3.26<br>(0.001)** | 12.66<br>(0.001)*** | 2.94<br>(0.004)** | 7.78<br>(0.007)** | 7.44<br>(0.008)** |
| FICA | 51.1 ± 9.8 | 40.3 ± 9.8 | 42.2 ± 9.1 | 38.4 ± 10.1 | 5.55<br>($<0.001$)*** | 30.74<br>($<0.001$)*** | 1.74<br>(0.087) | 1.64<br>(0.205) | 1.07<br>(0.304) |
| *Language* |  |  |  |  |  |  |  |  |  |
| PV | 56.9 ± 7.9 | 52.3 ± 6.9 | 53.5 ± 6.5 | 51.2 ± 7.2 | 3.16<br>(0.002)** | 7.76<br>(0.006)** | 1.45<br>(0.151) | 1.21<br>(0.276) | 1.28<br>(0.262) |
| *Processing speed* |  |  |  |  |  |  |  |  |  |
| PCPS | 53 ± 13 | 35.9 ± 13.3 | 38.1 ± 13.5 | 33.7 ± 12.9 | 6.49<br>($<0.001$)*** | 44.87<br>($<0.001$)*** | 1.44<br>(0.155) | 1.46<br>(0.232) | 1.53<br>(0.22) |
| *Temporal processing/Working memory* |  |  |  |  |  |  |  |  |  |
| RTCV | 0.15 ± 0.07 | 0.23 ± 0.14 | 0.20 ± 0.12 | 0.27 ± 0.16 | -3.31<br>(0.001)** | 14.34<br>($<0.001$)*** | 2.2<br>(0.031)* | 2.98<br>(0.089) | 2.36<br>(0.129) |

<sup>a</sup>ANCOVA covariates = Age, Gender, and Education<sup>b</sup>ANCOVA covariates = Age, Gender, Education, and Disease Duration
Presented as t-value (p-value) and F-value (p-value). \* p<0.05; \*\* p<0.01; \*\*\*p<0.001

When comparing PDFOG+ and PDFOG–, uncorrected independent *t-tests* revealed significant differences in MoCA cognitive assessment (p = 0.044) and DCCS executive function/attention test (p = 0.004) as well as RTCV of 7s interval timing temporal processing/working memory test (p = 0.03), but no other cognitive measurements (Table 2 and Fig. 1). After adjusting for covariates of age, gender, and years of education in the ANCOVA model, significant differences between PDFOG+ and PDFOG– remained for MoCA cognitive function assessment (p = 0.03), and DCCS test of executive function and attention (p = 0.007; Table 2). In the next model, when adding disease duration to the covariate adjustments of age, gender, and years of education, significant differences were again observed between PDFOG+ and PDFOG– for MoCA cognitive function assessment (p = 0.03), and DCCS measure of executive function and attention (p = 0.008; Table 2). However, similar to a previous study, when adjusting for covariates of age, gender, years of education, and disease severity (mUPDRS) in our complex model, differences between PD groups could not reach statistical significance (Table S1) [10]. Furthermore, we used individual covariates and controlled for them in the analyses in order to show their contribution to cognitive functioning in both PD groups. Overall, our models showed significant differences between PDFOG+ and PDFOG– for MoCA cognitive function and DCCS executive function/attention tests (Table S1).

**Fig. 1.**
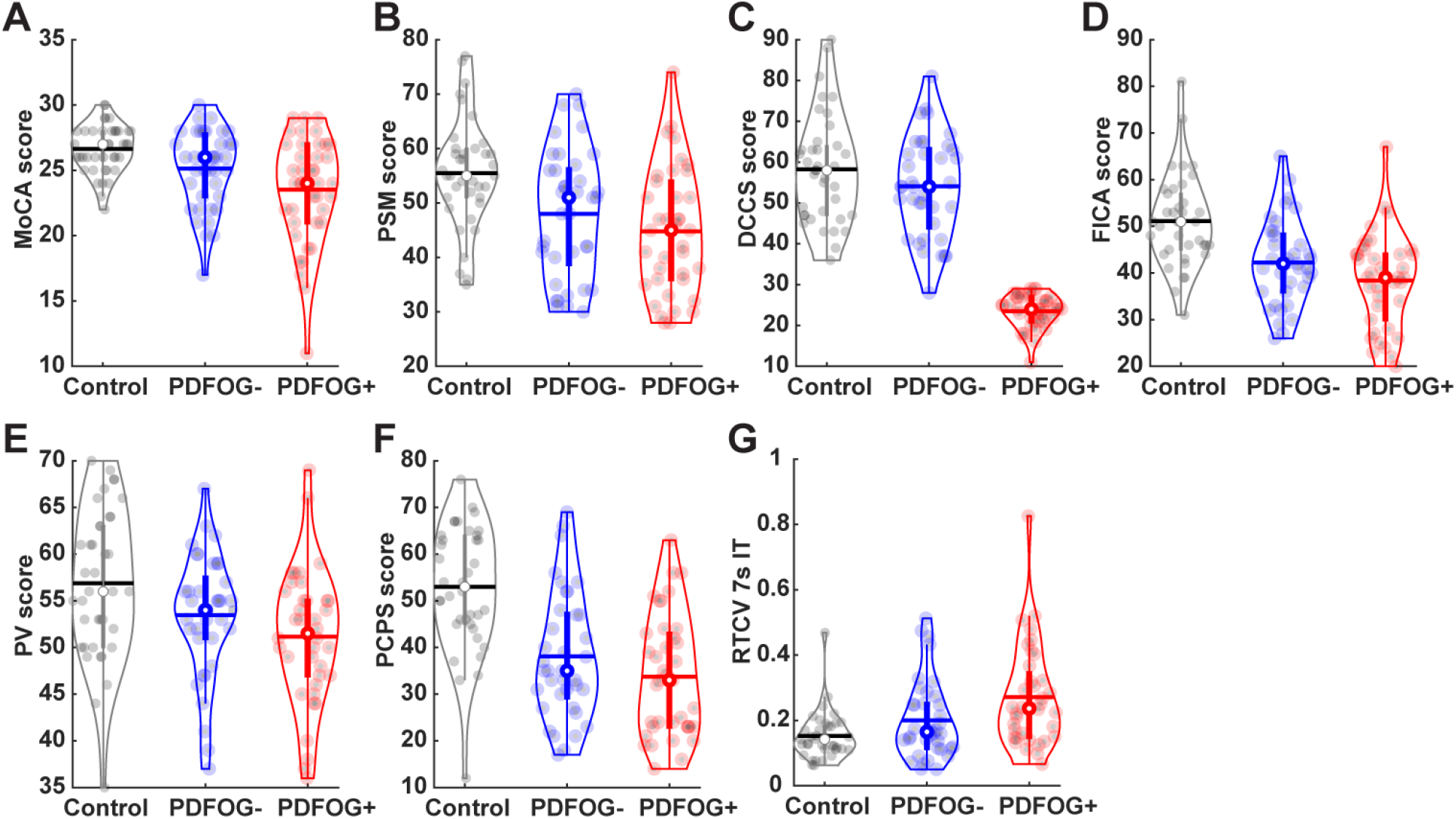
Violin plots display the cognitive measurements in healthy controls, PDFOG–, and PDFOG+. A) Montreal cognitive assessment (MoCA); B) picture sequence memory (PSM) test; C) dimensional change card sort (DCCS) test; D) flanker inhibitory control and attention (FICA) test; E) picture vocabulary (PV) test; F) pattern comparison processing speed (PCPS) test; G) Response Time Coefficient of Variation of 7-s interval timing task (RTCV). The horizontal lines and white circles in the violin plots represent the mean and median values, respectively.

### 3.3. Relationship between FOG severity and cognitive measurements

Correlation analyses, without controlling for covariates, suggested that FOG severity scores are significantly correlated with poorer cognitive scores for tests of cognitive function (MoCA, p = 0.015), episodic memory (PSM, p = 0.015), executive function/attention (DCCS, p = 0.014), and temporal processing/working memory performance (RTCV of 7s interval timing, p = 0.002) (Table 3 and Fig. S1). Even after controlling for age, gender, years of education, and disease duration, correlation analyses continued to show a significant relationship between FOG severity scores and poorer cognitive tests scores (Table 3).

**Table 3.**
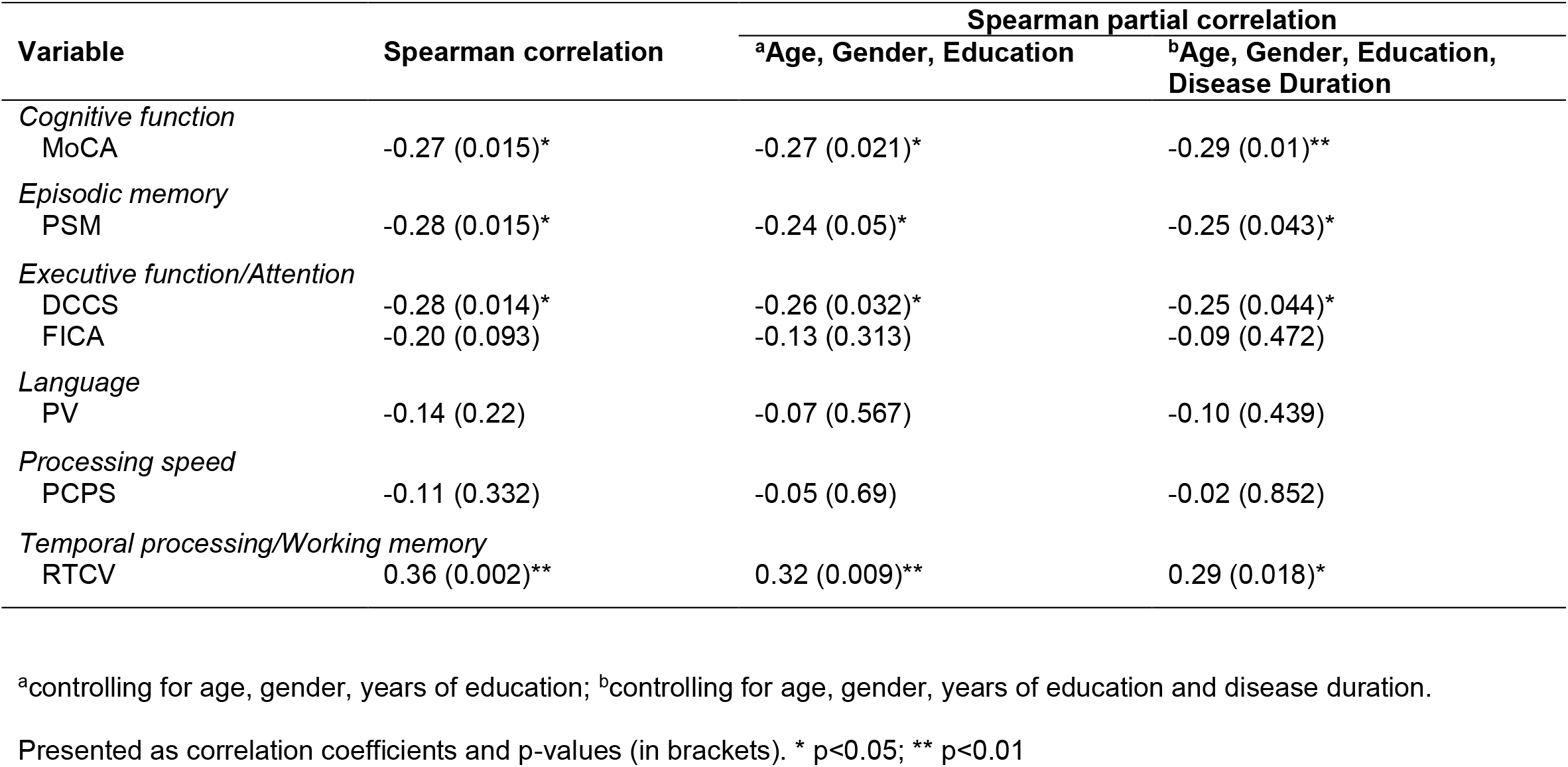
Correlations between FOG scores and cognitive measures.

Predicted responses of regression models showed significant positive relationships between disease severity (as determined by mUPDRS score) and FOG scores (R^2^ = 0.39, slope = 0.49, p < 0.001; Fig. 2) as well as between disease severity and MoCA score (R^2^ = 0.085, slope = -0.12, p = 0.03; Fig. 2). Predicted responses also indicated a significant relationship between disease duration and FOG score (R^2^ = 0.08, slope = 0.4, p = 0.01; Fig. S2); however, predicted responses showed no significant relationship between disease duration and cognitive function, as measured by MoCA score (R^2^ = 0.0001, slope = -0.003, p = 0.98; Fig. S2).

**Fig. 2.**
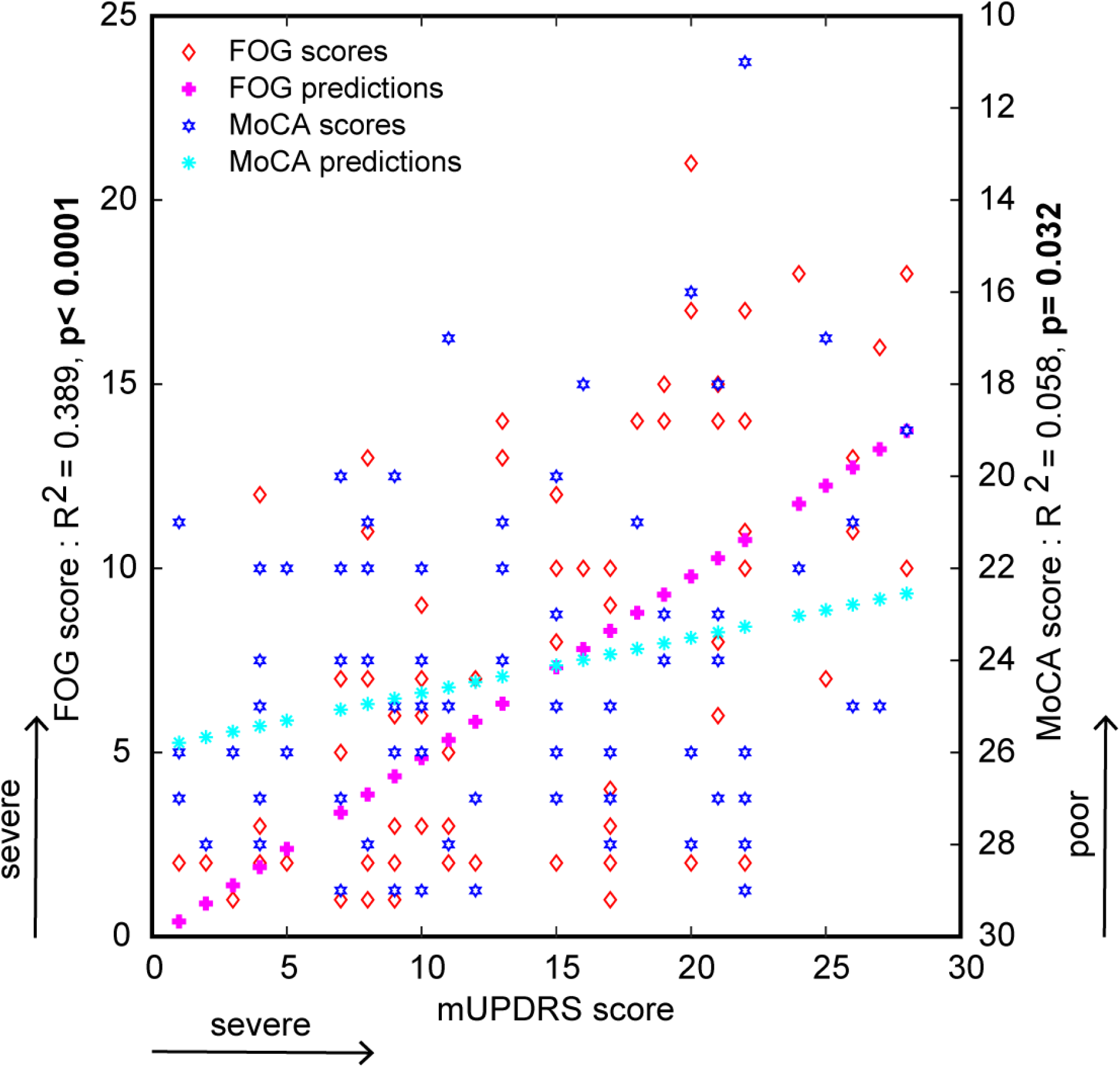
Linear regression analysis and predicted responses between disease severity scores and FOG severity and cognitive assessment (MoCA) scores. Bold p-values represent the significance.

Furthermore, mediation model showed a significant mediation effect of disease severity on the relationship between cognitive function (MoCA score) and FOG severity (FOG score) (ACME estimate = -0.22, p = 0.03) with significant direct effect of cognitive function (ADE estimate = - 0.32, p = 0.012) and total effect (p = 0.004). A second mediation model showed a significant mediation effect of cognitive function on the relationship between disease severity and FOG severity (ACME estimate = 0.039, p = 0.046) with significant direct effect of disease severity (ADE estimate = 0.45, p <0.0001) and total effect (p < 0.0001).

## 4. Discussion

FOG represents a significant problem for patients with PD. To better understand the relationships between FOG, disease severity and duration, and cognitive function, this study examined patients with PD both with and without FOG compared to controls, and found significant differences between controls and patients with PD for all cognitive domains. PDFOG+ and PDFOG– exhibited decreased performance in several cognitive tests, which persisted after adjusting for covariates that include disease duration, suggesting that FOG severity may be related to cognitive performance difficulties separate from disease duration. A meta-analysis detected a 71% FOG prevalence rate in patients with PD with disease durations ≥ 10 years [22], and patients with >12 years of disease duration developing severe cognitive impairments or PD dementia [23]. The disease duration for PD groups in the current study was not more than 6 years. Similar to our results, a previous study has also shown that PDFOG+ participants exhibit higher LEDD compared to PDFOG– and further suggested that high LEDD, cognitive impairment, and falls and balance problems can be independent determinants of FOG in patients with PD [24].

In general, the advancement of disease severity causes gait abnormalities and cognitive impairment in patients with PD, suggesting the presence of abnormal motor-cognitive networks. Moreover, our and prior studies have reported altered frontal theta and beta oscillations in the cortical-basal ganglia network during the performance of lower-limb motor and cognitive tasks in patients with movement disorders [5, 6, 12, 25, 26]. Typically, reduced frontal theta activity correlates with cognitive deficits in patients with PD [27], and similar abnormal activity has been observed at the time of lower-limb movement initiation in PDFOG+ [12], indicating the oscillatory relationship between gait and cognitive networks.

Differences in executive functioning and attention between PDFOG+ and PDFOG-in particular, lend interest to the role of these constructs in FOG. The current study demonstrated significant group-dependent differences in executive functioning that held true when controlling for disease duration, but not with disease severity. It has been suggested by others that release of inhibition associated with executive functioning deficits may be implicated in FOG [15]. Furthermore, our regression models suggest that FOG severity may be a predictor of more global cognitive impairment, and disease severity is correlated with both FOG and cognitive functioning (MoCA) scores. In contrast, disease duration predicted gait severity but not global cognitive impairment. This suggests that the relationships between FOG severity and cognitive impairment are not a direct result of disease duration. Mediation models suggest that cognitive function significantly mediates the relationship between disease severity and FOG severity, further suggesting that the role of cognition as a predictor of both FOG severity and overall PD severity needs to be further explored.

The effects of dopamine in cognitive and lower-limb motor control in patients with PD are not clear; several studies have shown no improvement in cognition and gait functions after levodopa medications [6, 12, 28]. In the current study, we assessed cognition in patients with PD with ‘on’ dopaminergic medication. Nonetheless, PDFOG+ showed differences in cognitive and FOG assessments compared to PDFOG–, suggesting an overload of the dopaminergic system in PDFOG+ may not improve or may worsen cognition and lower-limb movements, similar to levodopa-induced involuntary movements. Also, chronic levodopa treatment may lead to reduction in synaptic dopamine sensitivity, which may offer an explanation for cognitive and FOG severities in PDFOG+ and why these severities are not adequately improved by levodopa [29]. In our prior report, levodopa intake showed no improvement in frontal theta and beta oscillations in PDFOG+ during lower-limb pedaling motor task with cognitive load [12]; evidence suggests that frontal cognitive and motor functions are not affected by dopaminergic system in the advanced stage of parkinsonism. Therefore, it is critical to explore different pharmacological approaches to improving both cognitive and lower-limb motor functions in patients with advanced PD.

Furthermore, basal ganglia deep brain stimulation (DBS) therapeutic approaches have been implemented to improve cognitive and FOG severity in patients with PD. High-frequency basal ganglia DBS in the advanced PD stage influences only upper-limb motor performances; however, it affects neither lower-limb motor dysfunction (i.e. FOG severity) nor cognitive deficits [30]. Therefore, another target such as pedunculopontine nucleus (PPN) has been proposed to improve both cognitive and lower-limb motor functions in patients with advanced PD or PDFOG+, since pedunculopontine tegmental nucleus acts as an interface between the basal ganglia-cerebellum network, and PPB-DBS has the potential to influence cognitive and gait motor functions [31]. In addition, low-frequency DBS has been suggested to improve both cognition and FOG in patients with PD, since this approach can normalize frontal low-frequency and beta-band oscillations via cortico-basal ganglia networks [30, 32].

The current study had minor limitations which should be considered: our patients were assessed ‘on’ medication, and, therefore, our results should be interpreted accordingly compared to previous studies; we recruited only patients without DBS, so the effects of DBS on the relationship between cognitive and FOG severity could not be studied; and our patients did not perform gait or dual cognitive-gait task to measure quantitative information of FOG severity with and without cognitive load. Overall, the current study contributes to the body of evidence observing a link between cognitive function and FOG severity. Understanding the role of cognitive functioning in FOG may aid the development of therapeutic interventions that target cognitive domains such as executive functioning as a means of improving FOG qualities. Such approaches to therapeutic programming have shown initial success and promise toward improving the FOG behaviors in patients with a diagnosis of PD. Therapeutic approaches have successfully targeted explicit learning networks through cuing with the goals of movement recalibration and gait or FOG parameter improvements. Similarly, explicit cues have been used to improve behaviors compromised by executive functioning deficits. Cognitive training has shown initial success in reducing the FOG severity and offers promise that cognitive strategies may be harnessed to improve FOG behaviors in patients with a diagnosis of PD [13].

## Data Availability

Data will be available upon request.

## Acknowledgment

We would like to thank the participants who gave us their time to take part in this study.

## Supplementary Materials

**Fig. S1.**
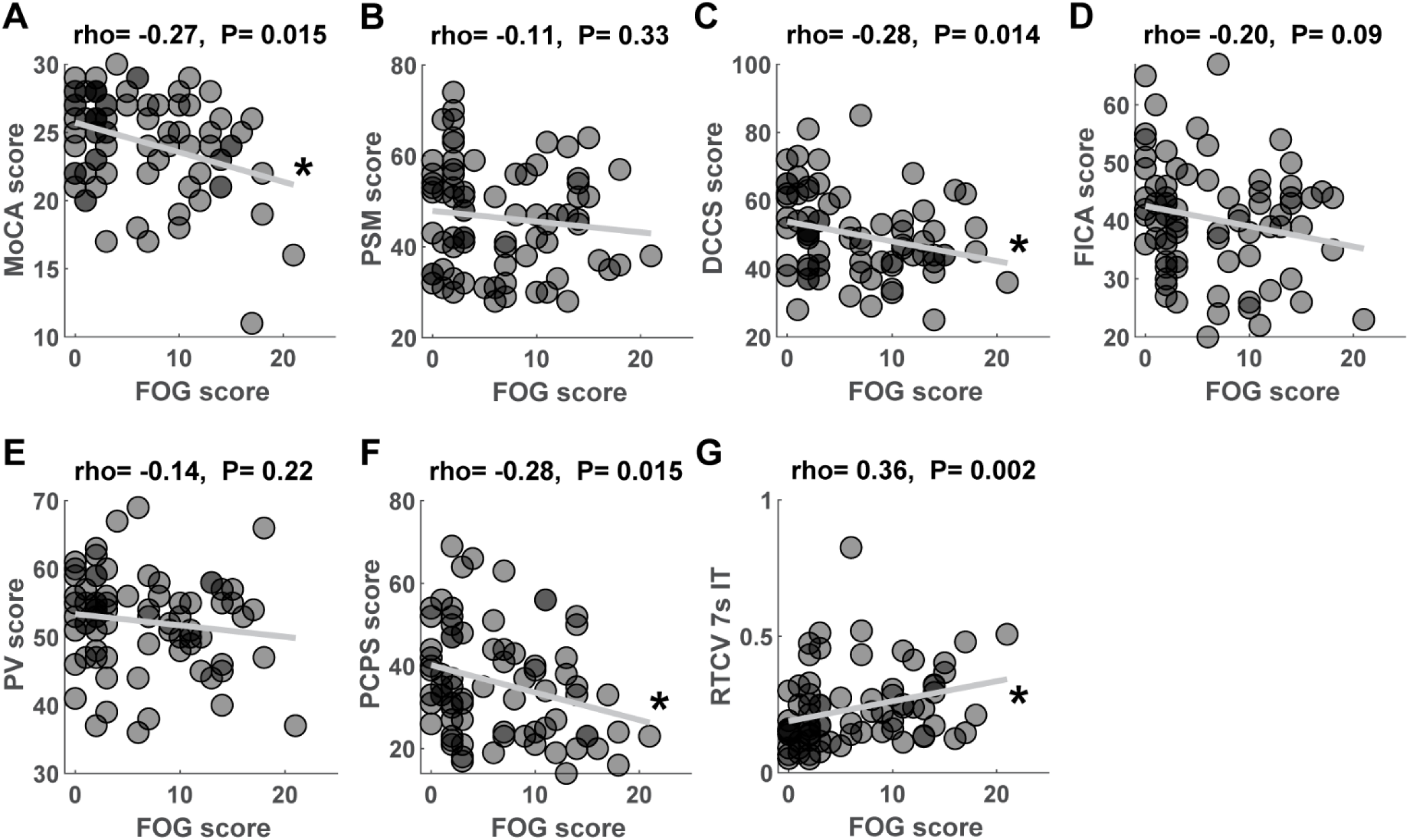
Spearman correlation plots between FOG scores and cognitive measurement scores (A-G) in PD patients. Higher FOG score denotes severe FOG, lower MoCA score (A) and NIH toolbox cognitive tests (B-F), and higher RTCV (G) denote poor cognitive control. * p<0.05 represents significant correlation.

**Fig. S2.**
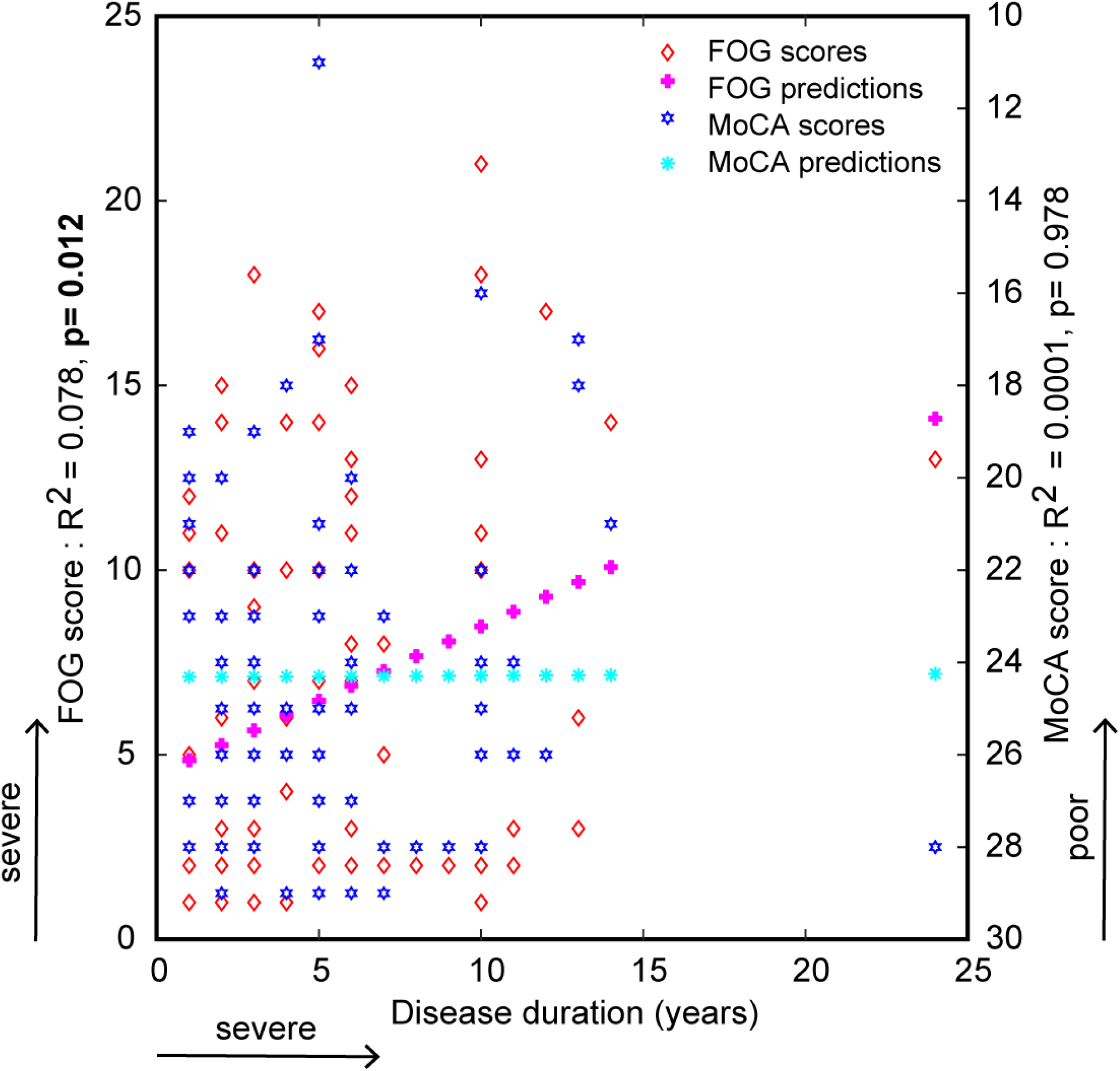
Linear regression analysis and predicted responses between disease duration and FOG severity and cognitive assessment (MoCA) scores. Bold p-value represents the significance.

**Table S1.** Cognitive Measurements between PDFOG+ and PDFOG- controlling for variables.

|  | Model 1 | Model 2 | Model 3 | Model 4 | Model 5 | Model 6 | Model 7 |
| --- | --- | --- | --- | --- | --- | --- | --- |
| Variable | Age | Gender | Years of Education | Disease Duration | Age, Gender, Education | Age, Gender, Education, Disease Duration | Age, Gender, Education, Disease Severity |
| *Cognitive function* |  |  |  |  |  |  |  |
| MoCA | 4.14 (0.045)* | 3.74 (0.057) | 5.22 (0.025)* | 4.3 (0.041)* | 4.55 (0.034)* | 4.7 (0.034)* | 0.68 (0.411) |
| *Episodic memory* |  |  |  |  |  |  |  |
| PSM | 0.99 (0.324) | 1.66 (0.202) | 0.68 (0.413) | 1.52 (0.222) | 0.049 (0.487) | 0.43 (0.515) | 0.58 (0.448) |
| *Executive function/Attention* |  |  |  |  |  |  |  |
| DCCS | 7.96 (0.006)** | 8.51 (0.005)** | 8.67 (0.004)** | 7.9 (0.006)** | 7.78 (0.007)** | 7.44 (0.008)** | 2.03 (0.159) |
| FICA | 2.45 (0.122) | 2.66 (0.107) | 2.4 (0.126) | 2.1 (0.152) | 1.64 (0.205) | 1.07 (0.304) | 0.18 (0.673) |
| *Language* |  |  |  |  |  |  |  |
| PV | 1.92 (0.17) | 1.81 (0.182) | 1.6 (0.21) | 2.06 (0.155) | 1.21 (0.276) | 1.28 (0.262) | 1.29 (0.259) |
| *Processing speed* |  |  |  |  |  |  |  |
| PCPS | 1.46 (0.231) | 1.9 (0.173) | 2.23 (0.14) | 2.03 (0.158) | 1.46 (0.232) | 1.53 (0.22) | 0.07 (0.79) |
| *Temporal processing/Working memory* |  |  |  |  |  |  |  |
| RTCV | 4.9 (0.03)* | 4.25 (0.043)* | 3.77 (0.056) | 3.99 (0.05)* | 2.98 (0.089) | 2.36 (0.129) | 0.50 (0.484) |

ANCOVAs for differences in cognition between PDFOG+ and PDFOG- controlling for variables. Model 1, adjusting for age; Model 2, adjusting for gender; Model 3, adjusting for years of education; Model 4, adjusting for disease duration; Model 5, adjusting for age, gender, years of education; Model 6, adjusting for all covariates; Model 7, adjusting for age, gender, years of education, and disease severity. Presented as F-value (p-value). \* p<0.05; \*\* p<0.01
MoCA: Montreal Cognitive Assessment
PSM: Picture Sequence Memory test of the NIHTB-CB
DCCS: Dimensional Change Card Sort test of the NIHTB-CB
FICA: Flanker Inhibitory Control and Attention test of the NIHTB-CB
PV: Picture Vocabulary test of the NIHTB-CB
PCPS: Pattern Comparison Processing Speed test of the NIHTB-CB
RTCV: Response Time Coefficient of Variation of 7-s interval timing task

